# Sample size bias in the empirical assessment of the acute risks associated with Daylight Saving Time transitions

**DOI:** 10.1101/2022.08.03.22278371

**Authors:** José María Martín-Olalla, Jorge Mira

## Abstract

The assessment of the acute impact of Daylight Saving Time (DST) transitions is a question of great interest for an understanding of the benefits and inconveniences of a practice that is now under public scrutiny in Europe and America.

Here we report a thorough analysis of a record of twelve well-known research studies that reported increased risks associated with DST transitions in health issues —acute myocardial infarction, ischemic strokes— and in societal issues —accidents, traffic accidents and fatal motor vehicle accidents—.

We found that a 5% increase of the risks suffices to understand the reported increased risks associated with the spring transition. Reported values above this threshold are impacted by the sample size of the study.

In the case of the autumn transition, no increase of the risks is found.

---

Sample size is the best-known study limitation of any research based on a sampled observation. Any finite sample size translates in a limited knowledge of the parent population that may have originated the observation. This limitation is essentially contained in the size of the confidence interval associated with the reported point estimate[1]. Researchers push hard to study large samples in the hope of a better determination of the effect. Nonetheless it is the case that the same phenomenon is analyzed by vividly different sample sized studies, which gives rise to a distribution of sampled results. Because every research study is analyzed on a solo basis, it is infrequent that the sample of reported point estimates is analyzed consistently in relation to the distribution of the sample sizes.

We bring here a comprehensive analysis of the research studies on the acute effects that Daylight Saving Time (DST) transitions may cause on public health and on societal issues. DST is the seasonal biannual changing of the clocks, a long-standing practice by which modern, extratropical societies —chiefly in America and Europe— adapt the phase of their daily rhythms to the early summer sunrises and to the late winter sunrises. The current criticism against the practice of DST focuses the disruption on human circadian rhythms it brings. The spring transition —when clocks are shifted forward— charges with the burden of the proof because the advance in the phase of human social rhythms is accompanied by a sleep deprivation which, eventually, may give rise to the increase in the incidence of acute diseases and accidents.[2, 3]

DST transitions set a natural experiment in which every individual participates. Yet when it comes to a research study that assesses the correlation between transition dates and societal issues, things are further limited: it is only one health or one societal issue that is analyzed in a limited region —a country, some region, or some hospital— and during a limited period of time —one year, a few years, one or two decades. All these factors impact the sample size of the study that can vary by some orders of magnitude, whereas the point estimate of the effect ranges from a factor 0.8 to a factor 1.9 — where no effect is associated with 1. Eventually, and in view of these results, governments in Europe and America are considering discontinuing this practice in the fear that its impact on human health may be more severe than previously thought[4], where, understandably, the largest point estimates may have been overrated and the role of the sample size underrated.

We analyze a set of thirteen well-known research studies that associated the incidence of acute myocardial infarction, ischemic strokes or traffic accidents with DST transition dates. Many of them were retrieved from review reports[2, 5]. They are frequently cited in review literature to support the discontinuity of DST practice[6, 7]. They are also cited in ex-post impact assessment reports from the European Parliament[4] or from the German Parliament[8] and in technical reports[9]. We show that the parent Poissonian population that may have originated the distribution of incidence ratios in this set of thirteen well-known research studies shows an increase of the risk around 5% in the spring transition. Any reported IR above this threshold is likely impacted by the sample size of the study.

## METHODS

The point estimate of the acute impact of DST transitions on human health is primordially assessed by the incidence ratio IR = *O/N* : the ratio of the observed counts *O* in a societal issue after a transition —also known as the study group— to the expected counts *N* in the same societal issue —the control group. The window of time to compute acute effects is the week after the transition. Research studies report either the week IR or the day IR in the first seven days following a transition, or both.

We included in our analysis research studies which, in addition to IR, reported the total counts in the control group *N* or this number could be deduced from the reading of the manuscript.

We found seven acute myocardial infarction (AMI) studies that met these requirements: six reported *O* in the first seven days following the spring and autumn transitions and deduced *N* from various estimates[10– 15]; one reported the IR for the first seven days after a transition[16] and *N* could be deduced from the total number of AMI in their catalogue. We also considered one ischemic stroke study[17] reporting IR in the first seven days after a transition.

We also considered a series of four accident-related studies: accidental deaths[18], traffic accidents[19], road accidents[20] and fatal motor vehicle accidents[21]. The first two studies reported *O* and *N* ; the last two studies reported IR and *N* could be deduced from their catalogue.

Finally, we considered one study on trauma admissions in Austria, Germany and Switzerland[22] which reported *O* and *N* for the week after and prior to the transition, excluding the transition date. This study did not report IR.

In addition to day IR and week IR we also analyzed a set of stratified IR associated with patient characteristics and co–diagnoses —among others: gender, age group, cholesterol in the first week.[11, 12, 16]. We also considered the breakdown by accident type —accident, motor-cycle accident, pedestrian, among others— in the trauma study[22].

Every of these studies were natural experiments with little design, in this way they are all comparable. The authors collected a dataset —such as the Swedish registry of AMIs[10], the National Highway Traffic Safety Administration registry of motor vehicle accidents[21] or the TraumaRegister DGU of the German Trauma Society— and crunched the numbers to get IR.

With the study group determined by the natural experiment, the control group —a contrafactual assessment of the events that would have been collected if the clock changing had not occurred— is the key parameter that gives rise to the reported IR. Studies differ in the way the control group is counted. Older, seminal studies take *N* as the number of events just before the transition,[19] or an average of event just before and well after the transition.[10] Modern approaches consider sophisticated models that take into account seasonal and societal variations.[20, 21] We do not distinguish the ones from the others and assume every author choice of *N* and, therefore, IR is a solid point estimate.

In our analysis we identify the total counts in the control group with the sample size of the study. Eventually the number comes from the population size of the region analyzed, the number of events —strokes or accidents— that this population produces every year, and the number of years in the catalogue. Table 1 summarizes the main characteristics of every study.

**Table 1.**
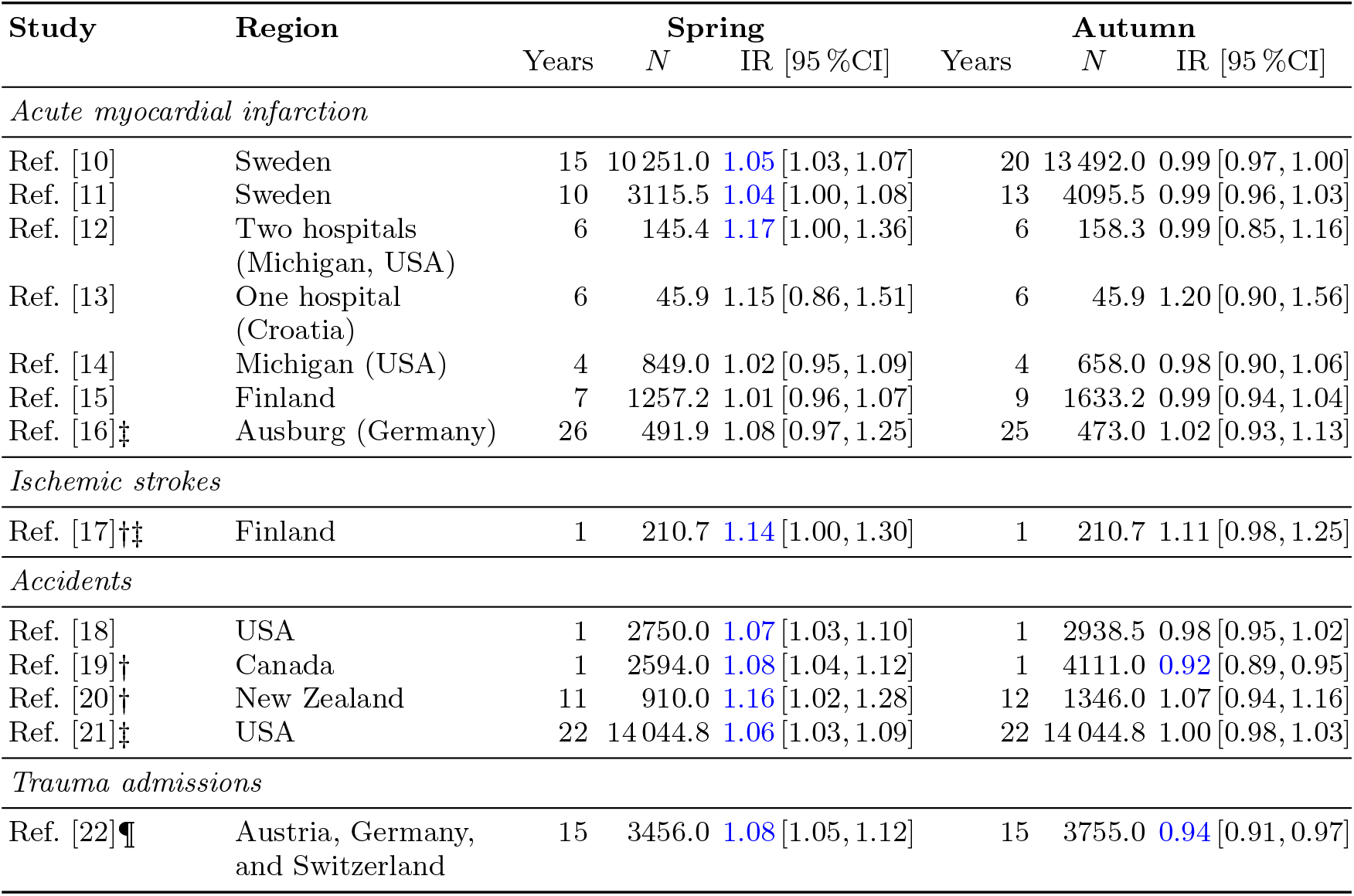
The sample size *N* —the total number of events in the control week, the expected total number of events from a model or the average number of events in a week— and the incidence ratio IR associated with the week after the spring transition and the autumn transition. Values were obtained for the week after a transition, except when marked with †, in which case the largest IR reported in the first week after a transition is shown. Studies marked ‡ with did not provide *N*, we deduced them from an average of the record. Studies marked with ¶ did not report IR and their 95%CI. Blue ink annotates *p*−values below the standard level of significance *α* = 0.05. According to their *N*, the reported values can be located inside the 95%CI for IR_*t*_ = 1.05 (spring), IR_*t*_ = 1 (autumn), see Figure 1 and Table 2.

Every study reported the 95% confidence interval (CI) associated with their point estimate of IR. Our goal in this meta-analysis is to test the distribution of reported IR and sample sizes *N* within the Poisson statistics. For this purpose we test parent populations with incidence ratio IR_*t*_ and retrieve exact Poissonian confidence intervals from the quantiles of the *χ*^2^ distribution for 2*N* · IR_*t*_ degrees of freedom (low bound) and 2 · (*N* · IR_*t*_ + 1) degrees of freedom (high bound)[23]. As IR_*t*_ is an adjustable parameter, we search for the value of IR_*t*_ whose 95%CI contains the 95% of the reported IR. In this way, the null hypothesis IR = IR_*t*_ sustains. In simple words, we will contextualize the difference between IR = 11 000*/*10 000 = 1.10 and IR = 110*/*100 = 1.10 within the Poisson statistics.

We do not include in our meta-analysis research studies on road traffic accidents where IR was not reported[24] (Finland) or *N* was unavailable[25] (Florida, USA). Likewise we do not consider research studies where the window of time was longer than one week[26] (United Kingdom) since they do not account for acute effects. Likewise we do not exclude any study on AMIs, ischemic strokes and accidents that reported *N* and IR to the best of our knowledge. We understand that the sample of thirteen research studies here analyzed which produced 64 day IRs, 10 week IRs, and 85 stratified IRs is representative of the literature on the field.

## RESULTS

Fig. 1 shows the reported IRs versus the sample size of the study for the spring transition (left) and the autumn transition (right). Generally speaking day IRs (intermediate ink) have smaller sample sizes than week IRs (darkest ink) and show a much larger variability.

**Figure 1.**
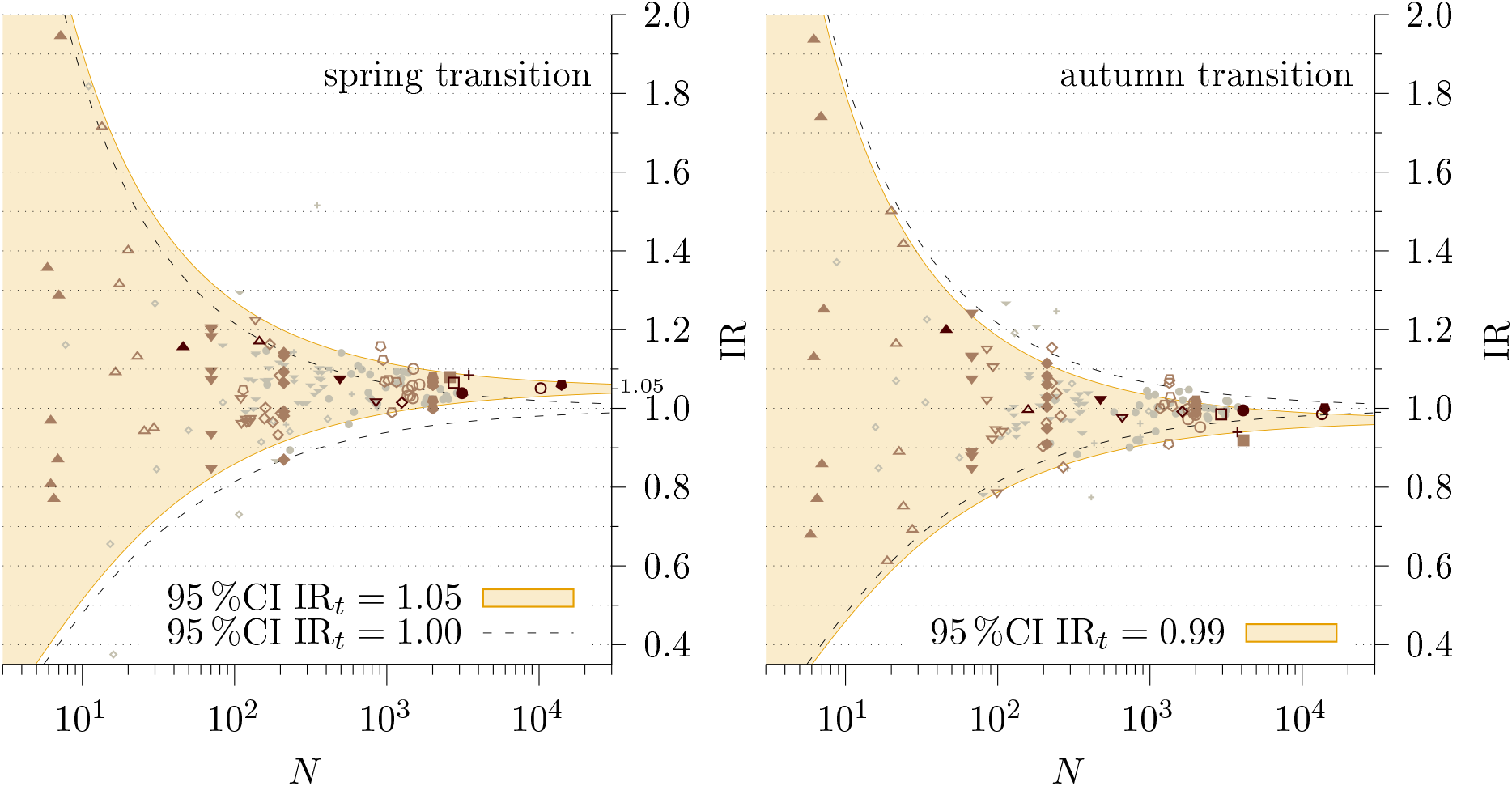
Scatter plot for stratified (lightest ink), day (intermediate ink) and week (darkest ink) IR associated with the spring transition (left panel) and the autumn transition (right panel). The lower and upper bounds of the 95% confidence interval for a Poissonian parent distribution with IR_*t*_ = 1.00 are shown by broken lines. The 95% confidence interval for a Poissonian parent distribution with IR_*t*_ = 1.05 (left) and IR_*t*_ = 0.99 (right) is shown in light shade background color. They roughly contains 95% of the observations in either panel, see Table 2. Legend: open circles[10], solid circles[11], open up triangles[12], solid up triangles[13], open down triangles[14], open diamonds[15], solid down triangles[16], solid diamonds[17], open squares[18], solid squares[19], open pentagons[20], solid pentagons[21], crosses[22]. See Table 2 for a breakdown of occurrences inside the 95% confidence intervals.

Fig. 1 left shows in broken lines the upper and lower bound of the 95%CI for 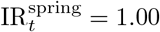, Table 2 summarizes the occurrences of reported IRs inside the 95%CI. In the first row of the table we see 59 (stratum, 75.6%), 47 (day, 73.4%) and 5 (week, 55.6%) reported IRs inside the 95%CI for 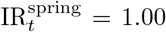. Therefore, the null hypothesis 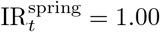 does not sustain at the standard level of significance (5%).

**Table 2.**
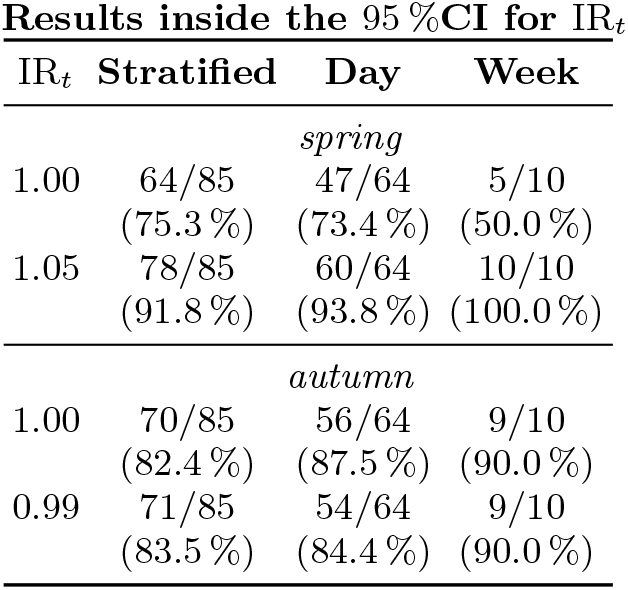
Occurrences of reported IRs inside the 95% confidence interval of a Poissonian parent distribution for selected IR_*t*_ (see Fig. 1 for a graphical representation). First the number of occurrences relative to the total number of observations; then, in parentheses, the percentage score. The null hypothesis IR = IR_*t*_ sustains at the standard level of significance for 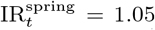. For the autumn transition both 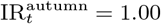 and 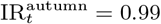 sustain similar scores.

| $IR_t$ | Stratified | Day | Week |
| --- | --- | --- | --- |
| *spring* |  |  |  |
| 1.00 | 64/85<br>(75.3 %) | 47/64<br>(73.4 %) | 5/10<br>(50.0 %) |
| 1.05 | 78/85<br>(91.8 %) | 60/64<br>(93.8 %) | 10/10<br>(100.0 %) |
| *autumn* |  |  |  |
| 1.00 | 70/85<br>(82.4 %) | 56/64<br>(87.5 %) | 9/10<br>(90.0 %) |
| 0.99 | 71/85<br>(83.5 %) | 54/64<br>(84.4 %) | 9/10<br>(90.0 %) |

In contrast the null hypothesis 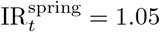 —whose 95%CI is noted by a light shade background color— yields the largest occurrences inside the CI: 72 (92.3%), 60 (93.8%) and 9 (100.0%) of the stratum, day and week observations (see second row in Table 2). Therefore the null sustains at the standard level of significance. A similar analysis for the autumn transition shows that the null hypothesis 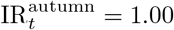 and 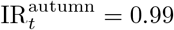 sustains with similar scores.

As a result we understand that for the spring transition the incidence ratio is close to 1.05 or 5% increase of the risks, while the autumn transition poses negligible small risks. Therefore, reported IRs above 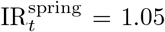 — that often raises the alarm in decision makers, the public opinion, and researchers— are likely impacted by the limitation that the sample size always brings.

There is only one clear outlier in the catalogue of reported IR: motorcycle accident admissions hits IR = 1.52 in the trauma study[22]. The authors explain that motor-cycle travels soars in Austria, Germany and Switzerland from March to April as many motorcyclists have summer license plates, which only operates from March to October.

## DISCUSSION

To understand the size of the spring effect we were able to estimate the relative sample standard deviation of the record in four studies[12, 14, 20, 21]. Figure 2 in [14] shows the residuals of the model on a daily basis. Visual perception suggests that the standard error of the model is at least some 20% of the daily AMIs, the largest reported IR is in line with this number. Table 2b in [20] provides the standard error of the model as 0.069 in the log_10_ space or 10^0.069^ = 1.17 in the linear space: their largest reported IR is in line with this number. On the other hand, Figure 1 in [21] shows the sample standard deviation of the weekly observation in two periods of eleven years each. From this we infer a relative sample standard deviation ∼ 8% in the week previous to the spring transition. Their reported IR is below this threshold. Finally, Table 1 in [12] contains *N*_*i*_ and *O*_*i*_ in the seven years of the record, from which we found a relative standard deviation of the sample ∼ 40% for one week. In contrast [12] reported a week IR = 1.17 or 17% increase in the week after the spring transition.

From these four studies —*N* ∈ (150, 14 000)— we understand that, while the spring transition shows a systematic increase of the risks, the size of the excursion is below the sample relative standard deviation of *O* every year. Therefore the impact of the transitions is below the myriad of things that populates the variability of the tested quantity, which explains why the assessment of the impact of the clock transitions on health issues is elusive.

## CONCLUDING REMARKS

We do not conclude from our analysis that DST transitions do not impact on public health. Instead, we bring attention to the fact that its impact might be as mild as previously thought. Decision makers and researchers should understand that the assessment of the IR is only the starting point of the balance of the risks associated with the practice. The risks of the practice should be balanced out against the risks of canceling a practice.

We bring the following recent example, US Senator Marco Rubio sponsored the Sunshine Protection Act of 2021 to “lock the clock”. Among other points, Rubio alleged that DST transitions caused 28 fatal motor vehicle accidents, a highlight from [21]. Much to the dismay of authors’ study and many others[6], Rubio vowed for locking the clock in the DST setting —that is, making DST the new perennial Standard Time— even though the study also showed that the risks of fatal accidents doubled after DST spring transition was advanced three weeks in the US after the Energy Policy Act[27]. Note that the Sunshine Protection Act of 2021 would effectively advance DST “spring transition” by ten weeks and delay DST “autumn transition” by eight weeks to cancel out both.

The thing to emphasize is that having locked the clock in the permanent Standard Time during the 20th century would have caused also a greater number of fatal accidents. In other words, the advance of clocks in spring —and, therefore, the delay of sunrise times— enforced by DST also helped to prevent risks. Had sunrise time occurred in the 40° latitude at 4.30am in summer, greater shares of population would have found comfortable to advance their daily activity relative to present day scores.

This choice would have translated into early winter activity, which is more prone to poor light conditions and, eventually, to greater accidental risks. Current research studies can assess the impact of the practicing transitions, as they set a natural experiment to analyze. On the contrary, we can only speculate with the impact of not having practiced DST transitions by analyzing the impact of past actions in the present day.[28]

## Data Availability

Data available from cited references.

## ACKNOWLEDGMENTS

The authors thank to Prof. Mercedes Conde Amboage from Universidade de Santiago de Compostela for fruitful discussion.

The authors declare no competing interest.

This project was not funded.

